# Efficacy of Povidone-Iodine Nasal And Oral Antiseptic Preparations Against Severe Acute Respiratory Syndrome-Coronavirus 2 (SARS-CoV-2)

**DOI:** 10.1101/2020.05.25.20110239

**Authors:** JS Pelletier, B Tessema, J Westover, S Frank, SM Brown, JA Capriotti

**Affiliations:** Ocean Ophthalmology Group (Miami, FL); ProHealth Physicians Ear, Nose and Throat (Farmington, CT); University of Connecticut, Department of Otolaryngology (Farmington, CT); The Institute for Antiviral Research at Utah State University (Logan, UT); Veloce BioPharma (Fort Lauderdale, FL)

## Abstract

**Introduction:** Severe Acute Respiratory Syndrome-Coronavirus 2 (SARS-CoV-2) is the pathogen responsible for the global pandemic of Coronavirus disease-2019 (COVID-19). From the first reported cases in December 2019, the virus has spread to over 4 million people worldwide. Human-to-human transmission occurs mainly through the aerosolization of respiratory droplets. Transmission also occurs through contact with contaminated surfaces and other fomites. Improved antisepsis of human and non-human surfaces has been identified as a key feature of transmission reduction. There are no previous studies of povidone-iodine (PVP-I) against SARS-CoV-2. This study evaluated nasal and oral antiseptic formulations of povidone-iodine (PVP-I) for virucidal activity against SARS-CoV-2. This is the first report on the efficacy of PVP-I against the virus that causes COVID-19.

**Methods:** PVP-I nasal antiseptic formulations and PVP-I oral rinse antiseptic formulations from 1-5% concentrations as well as controls were studied for virucidal efficacy against the SARS-CoV-2 virus. Test compounds were evaluated for ability to inactivate SARS-CoV-2 as measured in a virucidal assay. SARS-CoV-2 was exposed directly to the test compound for 60 seconds, compounds were then neutralized and surviving virus was quantified.

**Results:** All concentrations of nasal antiseptics and oral rinse antiseptics evaluated completely inactivated the SARS-CoV-2 virus.

**Conclusions:** Nasal and oral PVP-I antiseptic solutions are effective at inactivating the SARS-CoV-2 virus at a variety of concentrations after 60s exposure times. The formulations tested may help to reduce the transmission of SARS-CoV-2 if used for nasal decontamination, oral decontamination or surface decontamination in known or suspected cases of COVID-19.

## INTRODUCTION

The Severe Acute Respiratory Syndrome-Coronavirus 2 (SARS-CoV-2) virus has emerged as a new pathogen of the coronavirus family responsible for the Coronavirus Disease-2019 (COVID-19) pandemic. To date nearly 4 million cases have been confirmed. Since the beginning of the outbreak, ophthalmologists have played a pivotal role. In China they were among the first to recognize the emergence of the virus and were some of the earliest casualties among medical doctors. The virus is highly transmissible before, during and after the acute clinical phase of illness. Viral loads are high in the nasal cavity, nasopharynx and oropharynx^1^. Nasal goblet and ciliated cells within the respiratory epithelium have the highest expression of ACE2, the main receptor of COVID-19^2^. Viral shedding can be detected from nasal swabs before, during and after the onset of acute symptomatic disease including in seropositive antibody-converted convalescent cases^3,4^. Multiple reports have demonstrated that the nasal cavity, nasopharynx, and oropharynx are important routes of transmission^5,6^. Aerosol generating procedures can enhance this transmission via transit through these areas of high viral content, releasing aerosols that can remain in the air for up to 3 hours^7^. Transmission can occur in sub-clinical asymptomatic carriers, symptomatic infected carriers and convalescent seroconverted patients^8^. There is a growing need to develop processes to reduce virus transmission, as standard precautions including the donning of masks and gloves may not be sufficient. Early experience with COVID-19 outbreaks in hospital and healthcare settings have led frontline providers to suggest nasal and oral application of PVP-I as part of a transmission reduction plan^9^.

Nasal and oral antisepsis has been recommended as part of a comprehensive plan to reduce the likelihood of virus transmission by reducing the number of active aerosolized virus particles from the nasal passages and oral cavity^10,11^. Anesthesiologists, otolaryngologists, and oral surgeons have recommended specific protocols employing intranasal and intraoral PVP-I at dilute concentrations of 0.4-0.5% as a pre-procedure decontaminant. These groups also advocate that healthcare workers use intranasal and intraoral PVP-I - up to four times daily to reduce virus aerosolization. ^12^. The American Dental Association guidelines for minimizing risk of COVID-19 transmission advise use of PVP-I mouthwash prior to all procedures^13^.

In-vitro efficacy studies of PVP-I aqueous solutions have demonstrated concentration-dependent activity against a range of bacterial, fungal and protozoal pathogens^14,15^. Antiviral studies have confirmed activity against adenoviruses, rhinoviruses, coxsackieviruses and herpesviruses through presumed non-specific mechanisms^16^. Specific antiviral activity against influenza viruses involves receptor-mediated inhibition of hemagglutinin and neuraminidase pathways^17^. Interest in the use of PVP-I against coronaviruses was first reported in response to the SARS and MERS outbreaks in the past decade. Commercially available 10% PVP-I solutions have been tested against human coronaviruses HCoV 229E, HCV00C43, SARS and MERS^18^, though these commercial solutions are unsuitable for use in the nasal and oral cavities at commercially available concentrations. Homology with the current COVID-19 pathogen suggests that PVP-I might be effective, but there are no reported studies that have determined efficacy against SARS-CoV-2 for any PVP-I solutions^19,20^. No reported studies have evaluated povidone-iodine nasal antiseptics or oral rinse antiseptics specifically against the SARS-CoV-2 virus.

We report here the first anti-SARS-CoV-2 evaluation of a nasal antiseptic and an oral rinse antiseptic containing PVP-I which have been developed specifically for routine intranasal or oral use.

## METHODS

### Biosafety

All work with SARS-CoV-2 was conducted in biosafety level 3 (BSL-3) laboratories at The Institute for Antiviral Research at Utah State University (Logan, UT) following established standard operating procedures approved by the USU Biohazards Committee.

### Virus, media and cells

SARS-CoV-2, USA-WA1/2020 strain, virus stock was prepared prior to testing by growing in Vero 76 cells. Culture media for prepared stock (test media) was MEM with 2% fetal bovine serum (FBS) and 50 μg/mL gentamicin.

### Test compounds

Nasal antiseptic solutions and oral rinse antiseptic solutions consisting of aqueous povidone-iodine (PVP-I) as the sole active ingredient were supplied by Veloce BioPharma (Fort Lauderdale, FL). PVP-I concentrations of each solution as supplied and after 1:1 dilution are listed in **Table 1**.

**Table 1.** Virucidal efficacy of PVP-I antiseptic compounds against SARS-CoV-2 after a 60s incubation with virus at 22 ± 2°C.

| Test Product | PVP-I<br>Concentration (%)<br>After 1:1 Dilution | Incubation<br>Time | Virus<br>Titer<sup>a</sup> | LRV<sup>b</sup> |
| --- | --- | --- | --- | --- |
| PVP-I 5.0% Nasal Antiseptic | 2.5 | 60s | <0.67 | **4.63** |
| PVP-I 2.5% Nasal Antiseptic | 1.25 | 60s | 0.67 | **4.63** |
| PVP-I 1.0% Nasal Antiseptic | 0.50 | 60s | 0.67 | **4.63** |
| PVP-I 3.0% Oral Rinse Antiseptic | 1.5 | 60s | <0.67 | **4.63** |
| PVP-I 1.5% Oral Rinse Antiseptic | 0.75 | 60s | <0.67 | **4.63** |
| PVP-I 1.0% Oral Rinse Antiseptic | 0.5 | 60s | <0.67 | **4.63** |
| Ethanol 70% | na | 60s | <0.67 | **4.63** |
| Virus Control | **na** | **60s** | **5.3** | **na** |

<sup>a</sup> Log<sub>10</sub> CCID<sub>50</sub> of virus per 0.1 mL. The assay lower limit of detection is 0.67 Log<sub>10</sub> CCID<sub>50</sub>/0.1 mL.
<sup>b</sup> LRV (log reduction value) is the reduction of virus compared to the virus control

### Virucidal Assay

The test compounds were mixed directly with virus solution so that the final concentration was 50% of each individual test compound and 50% virus solution. A single concentration was tested in triplicate. Test media without virus was added to two tubes of the compounds to serve as toxicity and neutralization controls. Ethanol (70%) was tested in parallel as a positive control and water only as a virus control. The test solutions and virus were incubated at room temperature (22 ± 2°C) for 1 minute. The solution was then neutralized by a 1/10 dilution in MEM 2% FBS, 50 μg/mL gentamicin.

### Virus Quantification

Surviving virus from each sample was quantified by standard end-point dilution assay. Briefly, the neutralized samples were pooled and serially diluted using eight log dilutions in test medium. Then 100 μL of each dilution was plated into quadruplicate wells of 96-well plates containing 80-90% confluent Vero 76 cells. The toxicity controls were added to an additional 4 wells of Vero 76 cells and 2 of those wells at each dilution were infected with virus to serve as neutralization controls, ensuring that residual sample in the titer assay plate did not inhibit growth and detection of surviving virus. Plates were incubated at 37 ± 2°C with 5% CO_2_ for 5 days. Each well was then scored for presence or absence of infectious virus. The titers were measured using a standard endpoint dilution 50% cell culture infectious dose (CCID_50_) assay calculated using the Reed-Muench (1948) equation and the log reduction value (LRV) of each compound compared to the negative (water) control was calculated.

## RESULTS

Virus titers and LRV of SARS-CoV-2 after incubation with each of the nasal and oral antiseptics evaluated were effective at reducing >4 log_10_ CCID_50_ infectious virus, from 5.3 log_10_ CCID_50_/0.1 mL to 1 log_10_ CCID_50_/0.1 mL or less. No cytotoxicity was observed in any of the test wells. Positive control and neutralization controls performed as expected.

## DISCUSSION

Reopening of non-urgent clinical care environments and the re-commencement of elective surgical procedures in ophthalmology, otolaryngology and other specialties must be accompanied by attempts to reduce the likelihood of virus transmission. Current approaches to minimize COVID-19 transmission are anchored by three common strategies. First, the routine and widespread use of personal protection equipment (PPE) including masks forms a physical barrier to transmission ^21^. Second, the frequent and thorough disinfection of hands, surfaces and fomites which is important to mechanically remove and chemically inactivate shed virus particles and prevent their translocation to new hosts. Finally, nasal and oral decontamination with PVP-I is recommended to reduce the amount of virus particle aerosolization before it reaches barriers, surfaces and fomites^22,23,24,25^.

The challenge in nasal antisepsis is to find effective topical preparations which are safe to administer. Ethanol, for example, is known to be an effective virucidal agent but cannot be safely used in the nose^26,27,28^. PVP-I solutions are commonly used in health care settings as skin antiseptics, though most are contraindicated for intranasal use as they can decrease the ciliary beat frequency (CBF) at commercially available concentrations. Dilute concentrations below 1.25% do not have an inhibitory effect on ciliary beat frequency (CBF)^29^. They are well established in ophthalmology and commonly employed for preparation of the ocular surface and ocular adnexa. PVP-I concentrations of 2.5% and above are toxic to nasal mucosa^30^, upper airway respiratory cells^31^ and ciliated epithelia^32^. Despite their toxicity at higher concentrations, aqueous PVP-I solutions have been demonstrated to have concentration and contact-time dependent efficacy against a wide range of organisms^33^. We report here the first studies of PVP-I against SARS-CoV-2 in a virucidal assay. We also report the first and only anti-SARS-CoV-2 evaluation of nasal and oral antiseptics containing PVP-I preparations developed for safe, routine intranasal and intraoral use.

## CONCLUSION

The SARS-CoV-2 pandemic has reinforced the need for diligent attention to infection control, especially in ophthalmology and other outpatient healthcare related settings. Strict adherence to the use of physical barriers, spatial separation, and PPE are important aspects of any control program. In addition, chemical antisepsis remains a critical tool in the decontamination of fomites and surfaces, including surfaces found on patients and healthcare workers themselves. The nasal cavities, nasopharynx, oral cavity, and oropharynx of infected individuals all demonstrate high viral loads of SARS-CoV-2 and are principal reservoirs for the virus. There is a growing interest for decontamination of these areas in patients and healthcare workers to prevent virus transmission. PVP-I is of primary interest due to its ability to inactivate a broad range of pathogens, lack of microbial resistance, and long history of clinical use. The antimicrobial efficacy of PVP-I is highly dependent on the organism being eradicated, the PVP-I concentration and the antiseptic contact time. Though PVP-I has been shown to be a potent and broad virucidal agent active against even other members of the coronavirus family, specific activity against the SARS-CoV-2 virus had not been previously reported. The data reported here demonstrate the in vitro efficacy of PVP-I nasal and oral preparations specifically developed for use in the nasal passages, nasopharynx and oral cavities. Moreover, the antiseptics studied are rapidly virucidal at concentrations suitable for safe administration to the nasal and oral mucosa. Additional studies evaluating these formulations at contact times less than 60 seconds are being conducted to determine the range of exposure times over which virucidal activity is observed. The efficacy of these nasal and oral antiseptics against SARS-CoV-2 may support their use as additional hygiene measures in the COVID-19 outbreak. Additionally, the adoption of healthcare specific protocols utilizing PVP-I as an oral rinse and/or intranasally may be useful in decreasing viral burden in the outpatient setting.

## Data Availability

Data are presented in the manuscript.

